# Brain network changes after the first seizure: an insight into medication response?

**DOI:** 10.1101/2023.09.01.23294923

**Authors:** Mangor Pedersen, Heath R. Pardoe, Remika Mito, Moksh Sethi, David N. Vaughan, Patrick W. Carney, Graeme D. Jackson

## Abstract

After a first epileptic seizure, anti-seizure medications (ASM) can change the likelihood of having a further event. This prospective study aimed to quantify brain network changes associated with taking ASM monotherapy. We applied graph theoretical network analysis to longitudinal resting-state functional MRI (fMRI) data from 28 participants who had recently experienced their first seizure. Participants were imaged before and during long-term ASM therapy, with a mean inter-scan interval of 6.9 months. After commencing ASM, we observed an increase in the clustering coefficient and a decrease in network path length. Brain changes after ASM treatment were most prominent in the superior frontoparietal and inferior fronto-temporal regions. Participants with recurrent seizures display the most pronounced network changes after ASM treatment. This study shows changes in brain network function after ASM administration, particularly in participants with recurrent seizures. Larger studies that ideally include control cohorts are required to understand further the connection between ASM-related brain network changes and longer-term seizure status.

## Introduction

Anti-seizure medications (ASM) employ a variety of physiological mechanisms to decrease the frequency or intensity of seizures. Structural and functional MRI studies have shown brain changes associated with ASM treatment, either in response to successful seizure reduction or as an unintended side effect. For example, sodium valproate use is related to transient reductions in brain volume and cortical thickness in parietal and occipital cortices ^1,2^. fMRI studies have shown that ASM can increase task-based brain activation, particularly in association cortices ^3,4^ The fact that ASM reduces or eliminates abnormal electrographic activity suggests that they impact large-scale brain networks. These networks are complex systems of interconnected brain regions that work together to perform specific functions, such as sensory action, language and other higher-order cognitive functions. Epilepsy is characterized by disruption to brain networks, acutely during seizures but also chronically in the inter-ictal phase, which may contribute to developing epileptogenicity and the co-morbidities of epilepsy ^5^.

We aimed to study network effects in patients who experience their first seizure and commencing ASM treatment. To do so, we used fMRI data collected after the first seizure before ASM (pre-ASM) and after the start of long-term ASM treatment (post-ASM). Graph theoretical metrics were applied to quantify network organisation between the fMRI scans. Given the lack of prospective longitudinal studies on neuroimaging-based changes before and after commencing ASM, we conducted a whole-brain analysis of brain network changes following ASM administration.

## Methods

### Study cohort

We recruited 37 participants who were newly diagnosed with epilepsy and prescribed ASM treatment at two public hospitals, First Seizure Clinics in Melbourne, Australia. 9 patients did not participate in the follow-up MRI scan and were excluded from the study. The final cohort, therefore, consisted of 28 participants who were studied across two time points.

The cohort had a mean age of 32 ± 13 years, with 15 female and 13 male participants. The mean inter-scan interval was 6.9 ± 2.5 months, which coincided with clinical follow-up times. To summarise the clinical findings, 20/28 had focal epilepsy, 3/28 had generalised epilepsy, and 5/28 had an unknown epilepsy type. At the second scan, participants were taking a variety of ASMs, including Lamotrigine (14/28), Carbamazepine (6/28), Levetiracetam (4/28), Valproic Acid (3/28), Topiramate (1/28). In the 12 months following the first scan, 25% (7/28) participants experienced further seizures, whereas 68% (19/28) were seizure-free, and 7% (2/28) had unknown seizure status. These seizure freedom rates were similar to previous larger cohorts with new-onset epilepsy ^5^. Full clinical information is found in Table 1.

**Table 1:**
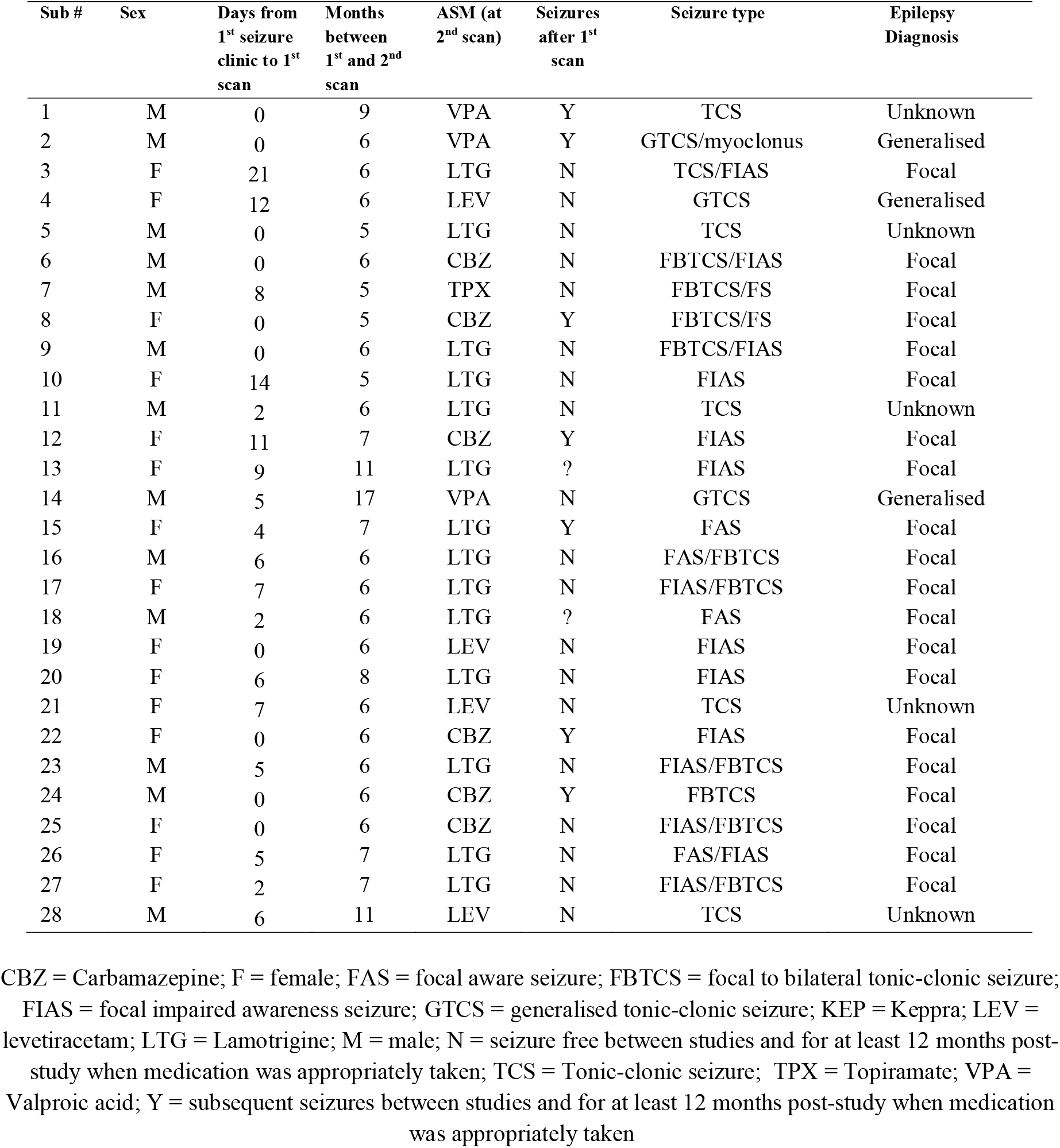
Participant information in this study.

The Austin Health Human Research Ethics Committee, Melbourne, Australia, approved this study, and subjects gave written consent to participate.

### fMRI network analysis

We pre-processed 15-minute long resting-state fMRI scans (pre-ASM and post-ASM) for each participant (TR = 3000 ms, TE = 30 ms and 3×3×3 mm voxel size) with fMRIPrep ^6^ (see supplementary materials 1, for complete fMRIprep methods). After fMRIPrep processing, the global signal, cerebrospinal fluid, white matter, and six movement parameters were regressed from the data (nine regressors in total). We observed no differences in head motion between scan sessions (paired t-test: *t*(27) = 0.70, *p* = 0.50) based on Framewise Displacement estimates ^7^. Then, the data was filtered between 0.01-0.1 Hz in FSL, comprising a nonlinear high pass and Gaussian linear lowpass filter. We parcellated the fMRI data using a Glasser parcellation mask with 180 symmetric brain regions (nodes) ^8^. Pearson *r* correlation was used to estimate functional connectivity between all nodes. We computed binary and proportionally thresholded networks between 20%-60% network density (i.e., the percentage of connections retained in the networks) with 5% increments using the Brain Connectivity Toolbox (https://sites.google.com/site/bctnet/). Test-retest of resting-state fMRI good, with a group, averaged correlation of *r* = 0.91 (supplementary materials 2).

Graph theoretical measures used in this study included *clustering coefficient, path length, betweenness centrality* and *normalized participation coefficient*. The *clustering coefficient* measures the number of interconnected local triangular nodes ^9^. The clustering coefficient, therefore, reflects the local segregation or clustering of nodes in a network. The *path length* of a network is estimated as the shortest possible path that connects two nodes. The path length is, therefore, a measure of network efficiency ^9^. The *betweenness centrality* measures brain hubs, reflecting the importance of a node in the network by calculating the shortest paths that pass through a node. Betweenness centrality values were normalised to have values between 0 and 1. Finally, the *participation coefficient* measures the amount of between-module connections and quantifies whether a node connects to its module or other modules ^10^. In our study, we performed a modularity decomposition by computing the Louvain community detection algorithm (in this context, modules are similar to resting-state networks) ^11^. We used a normalised version of the participation coefficient in this study ^12^, as this metric estimates the differences between real and randomised connectomes using Maslov and Sneppen network randomisations ^13^ to counter potential systematic effects between module size and node participation.

### Statistical analysis

The average of all network densities was used for statistical analysis. Within-subject paired-sample t-tests were used for statistical analysis. False discovery rate (FDR) was used to correct for multiple comparisons ^14^. To test the effects of seizure status, we also conducted a 2nd-level repeated measures ANOVA, where we quantified the main effects of time (network measures) and group (seizure-free versus not-seizure-free patient) interaction. Tukey’s tests were used to correct for multiple comparisons.

## Results

### Network differences between pre-ASM and post-ASM scans

After administering ASM, we found an increase in clustering coefficient (*t*(27) = 2.63, *p* = 0.02) and a decrease in path length (*t*(27) = -2.94, *p* = 0.01), averaged over all nodes. There was no global difference between pre-ASM and post-ASM for betweenness centrality (*t*(27) = 0.62, *p* = 0.54) or participation coefficient (*t*(27) = 1.31, *p* = 0.20). Results were corrected, *p*_*FDR*_ < 0.05 (Figure 1A). The clustering coefficient and path length are similar across subjects, whereas the participation coefficient is a distinct graph metric that correlates least with the other network measures (supplementary materials 3). Multiple linear regression analyses confirmed that these results were not confounded with age or time between scans (see supplementary materials 4)

**Figure 1:**
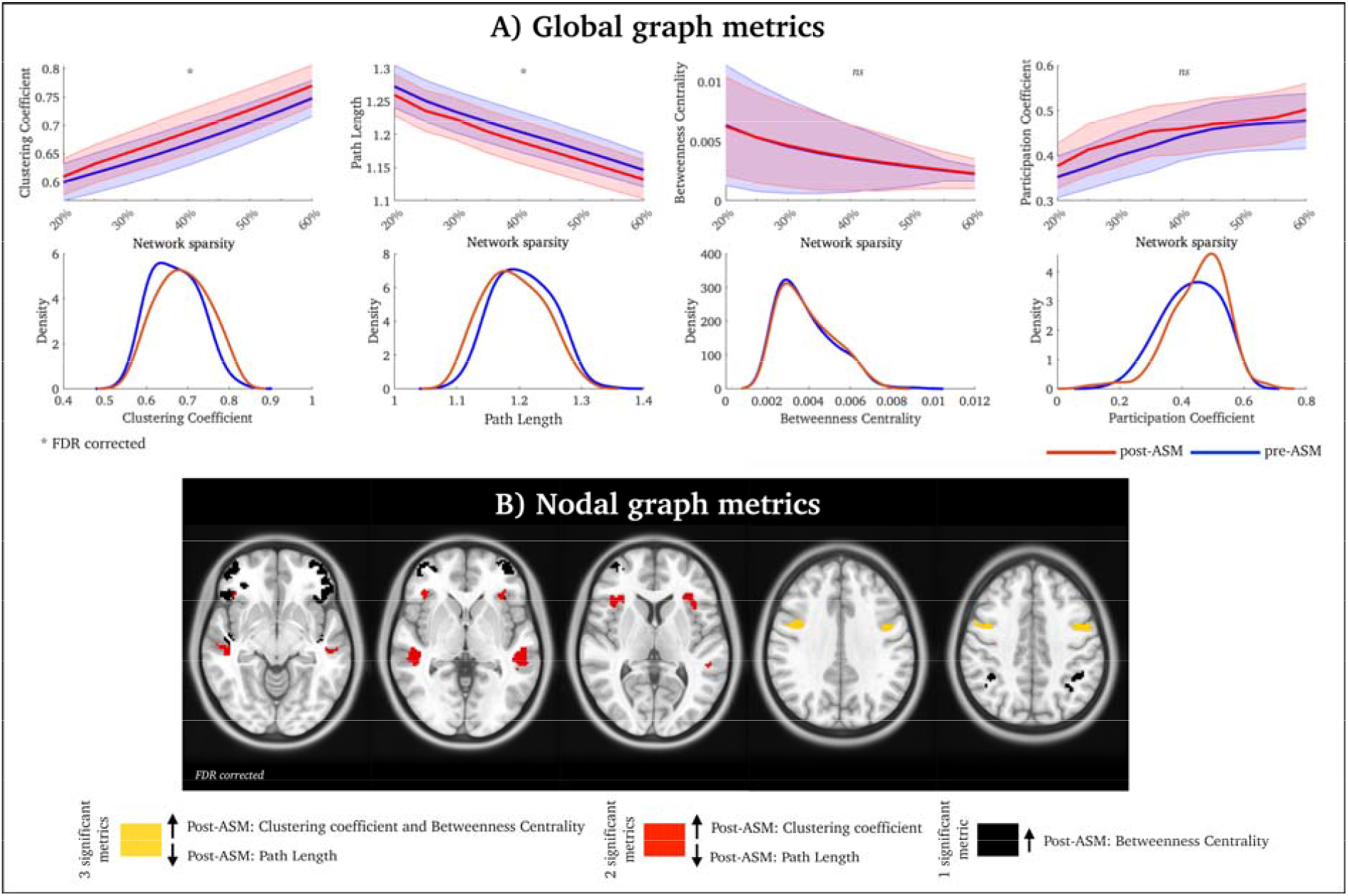
Network changes before versus after ASM therapy. A) Network topology results averaged over all brain nodes. Blue lines = pre-ASM; Red lines = post-ASM. The shaded error in the top row is the standard error. On the bottom row, we display the distribution. B) Significant nodes between pre-ASM and post-ASM scans (p_FDR_<0.05).

We also found that ASM administration resulted in changes in specific brain regions, including the superior frontal cortex (increased clustering coefficient and betweenness centrality as well as decreased path length, post-ASM – yellow nodes in Figure 1B) as well as the anterior insula and superior temporal gyrus (increased clustering coefficient and reduced path length, post-ASM – red nodes in Figure 1B). We also observed increased betweenness centrality post-ASM in the inferior parietal lobe and inferior frontal cortex – black nodes in Figure 1B. All nodal results were FDR corrected, *p*_*FDR*_ < 0.05.

### Increased clustering and decreased path length are associated with recurrent seizures

Next, we conducted an exploratory subgroup analysis quantifying network differences between people who experienced recurrent seizures (N=7) and people who remained seizure-free after the first scan (N=19). Two patients with unknown seizure status were excluded from this analysis.

Four repeated measures ANOVA was conducted to examine the effect of each global network on seizure status. The main effect of the clustering coefficient was significant, F(1,24) = 11.13, *p* = 0.003. There was also a significant interaction between the clustering coefficient and seizure status, F(1,24) = 4.86, *p* = 0.037. Tukey’s post hoc tests showed that not-seizure-free patients displayed a stronger clustering coefficient than seizure-free patients, *t*(24) = 3.24, *p* = 0.017.

The main effect of the path length was also significant, F(1,24) = 12.15, *p* = 0.002. There was also a significant interaction between the path length and seizure status, F(1,24) = 5.08, *p* = 0.034. Similar to above, not-seizure-free patients had a stronger path length than seizure-free patients, *t*(24) = 3.35, *p* = 0.013 (Tukey’s post hoc tests).

The betweenness centrality’s main effect was non-significant, F(1,24) = 2.68, *p* = 0.115. However, we observed a significant interaction between betweenness centrality and seizure status, F(1,24) = 4.43, *p* = 0.046. No significant post-hoc tests were detected.

The main effect of the participation coefficient was not significant, F(1,24) = 1.92, *p* = 0.183. No significant interaction existed between participation coefficient and seizure status, F(1,24) = 0.14, *p* = 0.714. There were no significant post-hoc tests.

See Figure 2 for global (Figure 2A) and nodal network results (Figure 2B).

**Figure 2:**
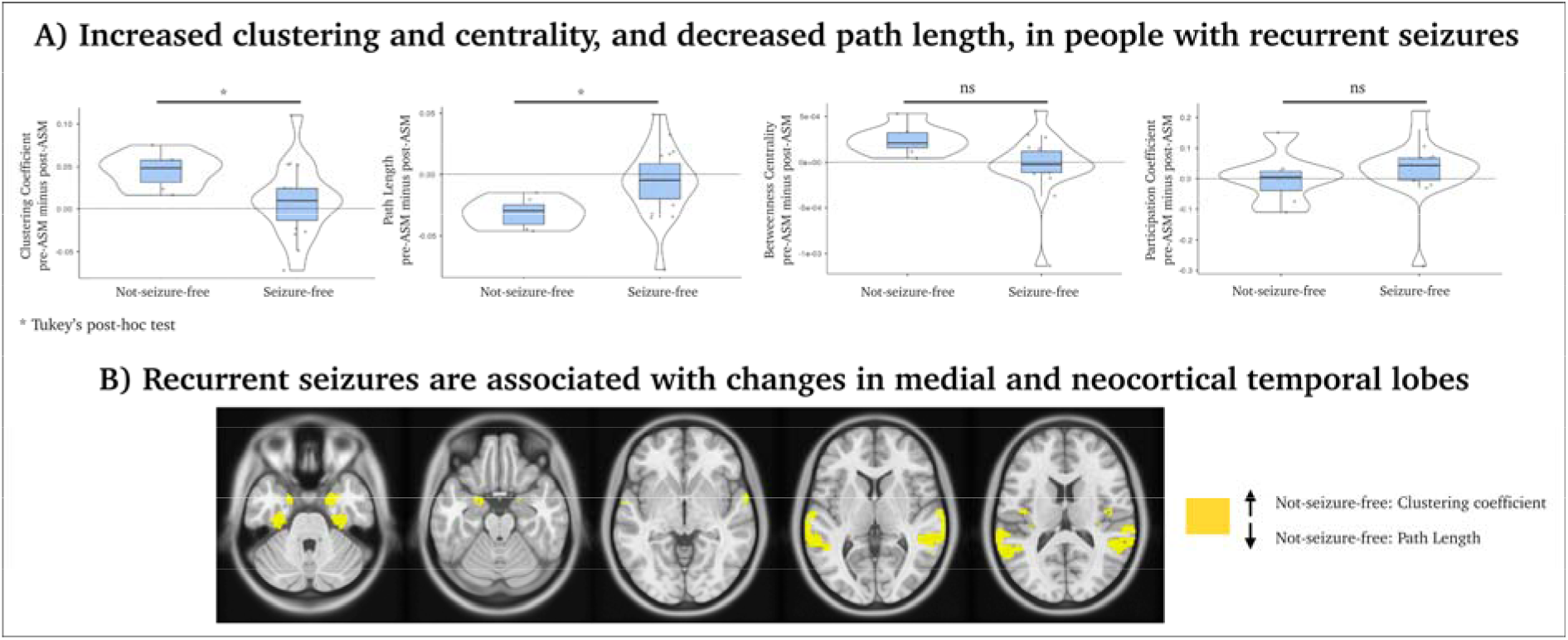
Comparison between seizure-free and non-seizure-free participants. A) Network topology results averaged over all brain nodes between people with recurrent seizures and no seizures after the first scan, averaged across all network sparsity thresholds. Each dot is a subject, the median is highlighted as a black line, and 95th Percentile intervals are shaded blue. B) Independent-samples t-test of network differences between seizures-free and not-seizure-free patients, p<0.01, uncorrected

### No network difference between Lamotrigine and other ASM

Next, we conducted a subgroup analysis between people on Lamotrigine (N=14) and other ASM (N=14). We observed no significant difference between Lamotrigine and other ASM on a whole-brain averaged and region-specific level in our four network metrics.

## Discussion

This proof-of-principle study observed brain network changes after people commenced ASM treatment, reflecting increased network clustering and efficiency (Figure 1). Our findings suggest that these changes are predominantly driven by patients who experience recurrent seizures after commencing ASM treatment, suggesting that these brain network changes may be a consequence of ongoing seizure activity. Although we still do not have consistent functional network markers of the epilepsies ^15^, studies suggest that recurrent seizures are associated with a greater clustering coefficient and reduced path length ^16^, as found amongst our patients with recurrent seizures. Several network models also hypothesise that increased clustering and decreased path length are markers of seizure topology ^17–21^. In addition, our study has shown that recurrent seizures are associated with changes in mesial and neocortical temporal lobes (Figure 2). This may indicate that changes in fMRI connectivity between scans are potential markers of recurrent seizures, and in our case, it may predict a failure to respond to ASMs. Further research on larger datasets will be crucial to improving our understanding of the dynamic changes in connectivity in varied brain regions in specific epilepsies, which might prove useful clinical tools in the future.

Patients with recurrent seizures explained most, but not all, of the variance in our data. This may explain the different brain patterns we observed in all patients (Figure 1), in contrast to the sub-analysis with seizure-free versus not-seizure-free patients (Figure 2). In line with previous reports, our findings may include network mechanisms involved in cognitive functions after the administration of ASM, including working memory and language (see Figure 2). Other studies assessing ASM effects on brain function have shown valproate increased fMRI signal in the frontoparietal area during a working memory task ^23^, and changes in precuneus activity were associated with ASM dosage in a verbal fluency task ^24^. These observations may provide insights into how ASMs may cause cognitive side effects or alternatively, given evidence of cognitive change prior to epilepsy diagnosis ^25^, ASM treatment may specifically impact on these already disrupted networks having a positive impact on seizure control.

A limitation of this study is that we did not have longitudinal data from healthy controls, so we cannot attribute normal versus abnormal network changes in the current study. Future studies may benefit from including control populations to compare the effects of different types of medication over time on people without a seizure history. Nevertheless, there is a great need for reliable biomarkers of treatment response in epilepsy ^26^. In a computational study, Woldman et al. ^22^ proposed that the recurrence of seizures may be associated with alterations in brain region connectivity, as found in our study, providing a plausible model for understanding the underlying mechanisms of ASM responses. Prospective studies, such as ours, play a crucial role in obtaining brain markers of treatment response. However, it is important to acknowledge the challenges associated with conducting such studies, including issues related to its longitudinal nature and the high dropout rates observed during follow-up scans (25% drop-out rate in our study). Our network findings may also be helpful for disorders beyond epilepsy to monitor brain changes in neurological and psychiatric disorders where pharmacological interventions are used.

## Data Availability

Anonymised data is available upon reasonable request to the authors.

## Acknowledgements

We thank Magdalena Kowalczyk, Donna Parker and Mira Semmelroch for recruiting participants for this study.

