## Supplementary Materials for "Brain network changes after the first seizure: an insight into medication response?"

**Supplementary Materials 1**

A T1-weighted (T1w) T1w image (TR = 1900 s, TE = 2500 ms and 0.9x0.9x0.9 mm voxel size) and two 15-minute long resting-state fMRI scans (pre-ASM and post-ASM) were completed for each participant (TR = 3000 ms, TE = 30 ms and 3x3x3 mm voxel size). Data preprocessing was performed using fMRIPrep ^20,^ , which is based on Nipype ^2^. T1w images were corrected for intensity non-uniformity with N4BiasFieldCorrection ^3^ in ANTs ^4^. The T1w-reference was then skull-stripped with a Nipype implementation of the antsBrainExtraction.sh workflow (from ANTs), using OASIS30ANTs as the target template. Brain tissue segmentation of cerebrospinal fluid, white matter and grey matter was performed on the brain-extracted T1w using fast in FSL (the FMRIB Software Library) ^5^. An anatomical T1w-reference map was computed after registration of the T1w images (after intensity non-uniformity correction) using mri_robust_template in FreeSurfer ^6^. Brain surfaces were also reconstructed using recon-all in FreeSurfer ^7^, and the brain mask estimated previously was refined with a custom variation of the method to reconcile ANTs-derived and FreeSurfer-derived segmentations of the cortical grey matter of Mindboggle ^8^. Volume-based spatial normalization to standard spaces (MNI152NLin6Asym, MNI152NLin2009cAsym) was performed through nonlinear registration with antsRegistration, using brain-extracted versions of both T1w reference and the T1w template. A deformation field to correct susceptibility distortions was estimated based on fMRIPrep's fieldmap-less approach. The deformation field results from co-registering the fMRI reference to the same-subject T1w-reference with its intensity inverted ^9,10^. Registration was performed with antsRegistration, and the process is regularized by constraining deformation to be nonzero only along the phase-encoding direction and modulated with an average field map template ^11^.

The following preprocessing was performed for each fMRI session (pre- and post-ASM). First, a reference volume and its skull-stripped version were generated using a custom methodology of fMRIPrep. Head-motion parameters for the fMRI reference (transformation matrices and six corresponding rotation and translation parameters) are estimated before spatiotemporal filtering using mcflirt in FSL ^12^. The calculated field map was aligned with rigid registration to the target echo-planar imaging reference run. The field coefficients were mapped onto the reference echo-planar imaging using the transform. The fMRI reference was then co-registered to the T1w reference using bbregister (FreeSurfer), which implements boundary-based registration ^13^. Co-registration was configured with six degrees of freedom. The signals are extracted within the cerebrospinal fluid, the white matter, and the whole-brain masks (i.e., the global signal). The fMRI time series were resampled into standard space, generating a preprocessed fMRI run in MNI152NLin6Asym space. First, a reference volume and its skull-stripped version were generated using a custom methodology of fMRIPrep. All resamplings can be performed with a single interpolation step by composing all the pertinent transformations (i.e. head-motion transform matrices, susceptibility distortion correction when available, and co-registrations to anatomical and output spaces). Gridded (volumetric) resamplings were performed using antsApplyTransforms (ANTs), configured with Lanczos interpolation to minimize the smoothing effects of other kernels ^14^. Non-gridded (surface) resamplings were performed using mri_vol2surf in FreeSurfer.

**Supplementary Materials 2:**

*Test-retest analysis*

A Pearson correlation range between pre- and post-ASM from *r* = 0.36 to *r* = 0.86 on an individual level. After averaging the fMRI connectivity edge values across all subjects, Pearson’s *r* was 0.91. This finding suggests a good reproducibility between fMRI scans across subjects (Figure 3).

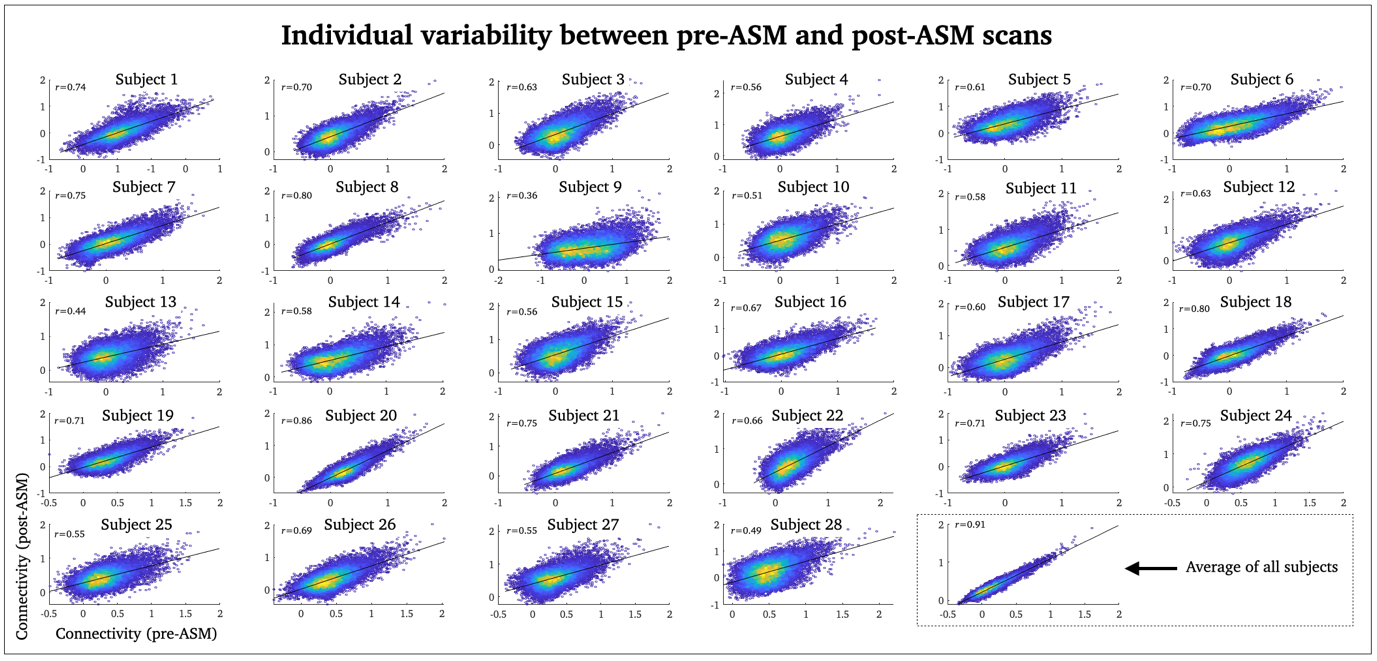

***Figure 3:*** *In these scatterplots, the data points represent edge-wise Fisher’s z-scored correlation values from each patient’s 180 x 180 connectivity matrix. The first scan (pre-ASM) is on the x-axis, and the second scan (post-ASM) is on the y-axis.*

**Supplementary Materials 3:**

Similarity (Pearson’s r) between graph metrics used in this study, averaged across subjects and scans.

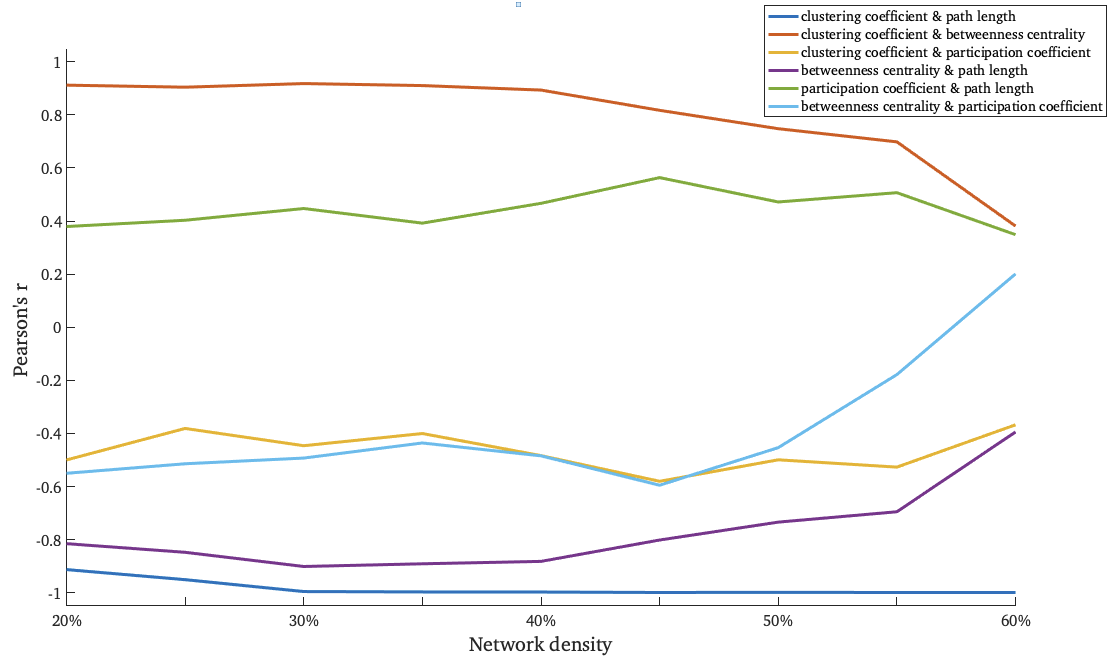

**Supplementary Materials 4:**

Scatter plots and Pearson’s correlation values between differences in network metrics between scans (pre-ASM and post-ASM), and people’s age at the first scan (top row) and months between the pre-ASM and post-ASM scan (bottom row). No correlation was statistically significant at *p* < 0.05.

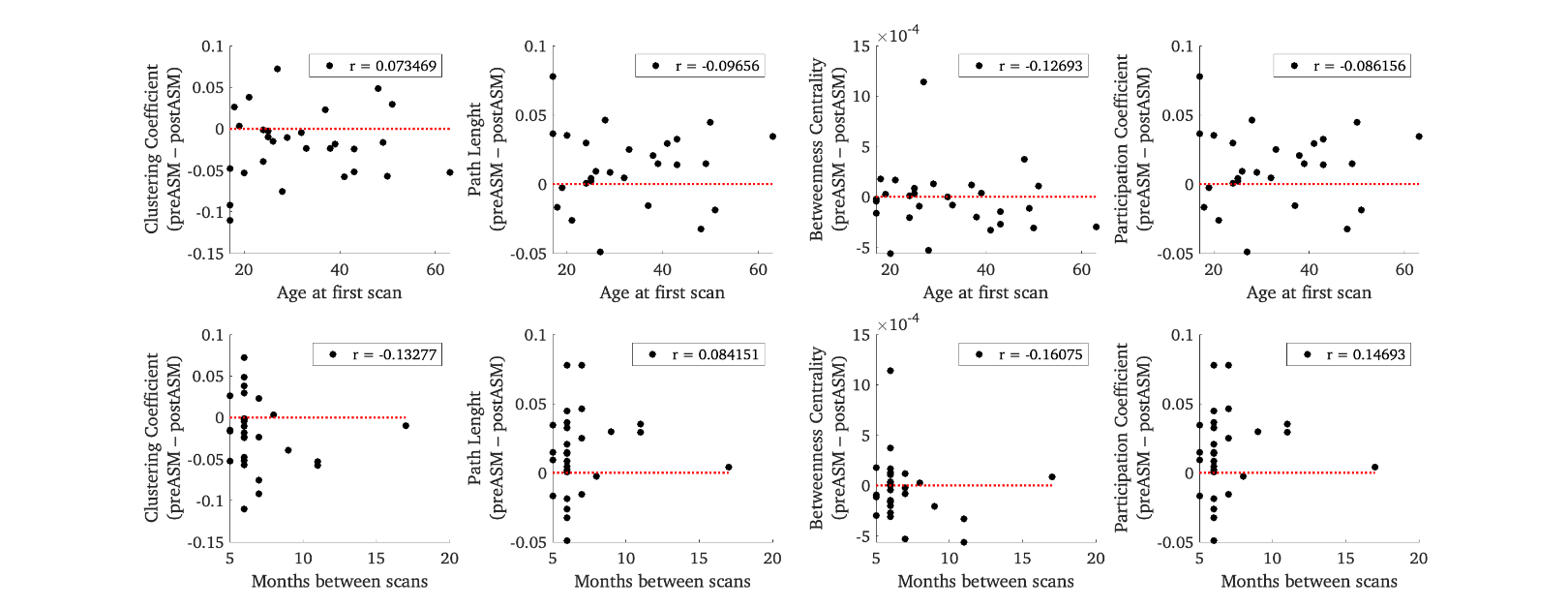

We also conducted 4 linear regression models (difference between pre-ASM and post-ASM for the 4 network metrics), with age and months between scans as covariates. No significant regressions were seen for the clustering coefficient (*R*^2^ = .02, F(2, 25) = 0.26, p = .772), path length (*R*^2^ = .01, F(2, 25) = 0.15, p = .863), betweenness centrality (*R*^2^ = .10, F(2, 25) = 1.47, p = .249) or participation coefficient (*R*^2^ = .03, F(2, 25) = 0.35, p = .725).
